# Adverse outcomes in SARS-CoV-2 infections are associated with a combination of variant genotypes at two loci in the *APOL1* gene: a UK Biobank study

**DOI:** 10.1101/2021.11.02.21265755

**Authors:** Walt Adamson, Harry Noyes, Anneli Cooper, Georgia Beckett-Hill, John Ogunsola, Rulan S. Parekh, Annette MacLeod

**Affiliations:** Wellcome Centre for Integrative Parasitology (WCIP). Institute of Biodiversity, Animal Health, and Comparative Medicine, University of Glasgow, Glasgow, UK; Institute of Biodiversity, Animal Health and Comparative Medicine (IBAHCM), University of Glasgow, Glasgow, UK; Centre for Genomic Research, University of Liverpool, Liverpool, UK; TrypanoGEN Research Group; Women’s College Hospital, Hospital for Sick Children and University of Toronto, Ontario, Canada

## Abstract

Risk of hospitalisation or death from COVID-19 in the UK is disproportionately high in people of African ancestry. Two *APOL1* haplotypes (G1 and G2) found at high frequency only in populations of African descent are associated with increased risk of non-communicable and infectious diseases. Here, we test the hypothesis that adverse COVID-19 outcomes are also associated with these *APOL1* high-risk variants.

Within 9,433 individuals with African ancestry in the UK Biobank, there were 172 hospitalisations and 47 deaths attributed to COVID-19 as of December 2021. We examined *APOL1* genotypes for association with hospitalisation and death while controlling for risk factors previously associated with poor COVID-19 outcomes.

We identified an association between carriage of two *APOL1* high-risk variants and death from COVID-19 (OR=2.7, 95% CI: 1. 2-6.4). Stratified by genotype, those with G1/G2 had a higher odds of COVID-19 hospitalisation (OR=2.1, 95% CI: 1.1-3.8) and death (OR=5.9, 95% CI: 2.2-15.3) than G0/G0. There was no significant association detected in carriers of G1/G1 and G2/G2.

These data suggest that the APOL1 G1/G2 genotype contributes to the increased rates of hospitalisation and mortality from COVID-19 in people of African ancestry, and could help to identify those at higher risk of severe COVID-19. This is especially relevant to geographical regions where *APOL1* G1 and G2 high-risk variants are common, such as West and Central Africa and their diaspora.

## Introduction

The COVID-19 global pandemic was first identified in Wuhan, China in December 2019^1^. By the beginning of 2022, the disease (caused by the SARS-CoV-2 virus), had claimed the lives of nearly 6 million people world-wide^2^. In the UK by December 2021 there were 10 7 million PCR-confirmed cases and 145,000 deaths^2^, with the risk of hospitalisation or death from COVID-19 being disproportionately higher in people of African ancestry, for reasons not fully understood^3^.

People of recent African origin are also disproportionately affected by other conditions, such as chronic kidney disease, HIV-associated nephropathy, sickle cell nephropathy, preeclampsia, and sepsis^4,5^. Excess risk of these diseases has been, in part, attributed to carriage of variants at two independent loci of the apolipoprotein L1 (*APOL1*) gene^6^. These variants, termed G1 (encoding S342G and I384M) and G2 (encoding deletion of N388 and Y389) are found within the same protein domain and are present on separate African-derived haplotypes^6^, which are in complete negative linkage disequilibrium. Haplotypes containing both G1 and G2 variants are theoretically possible but due to their close physical proximity have not yet been reported. Non-variant alleles at these positions are termed G0. This haplotype arrangement gives rise to nine possible genotypes with respect to *APOL1* G1 and G2, of which six have been reported (Supplementary Table 1). G1 and G2 variants arose to high frequency under strong selection within the last 10,000 years^6^ and are common in the African diaspora (with estimated allele frequencies in African Americans of 21% and 13%, respectively^6^) but are either absent or occur at very low frequency in non-African-derived populations.

**Table 1:** Risk of hospitalisation or death due to COVID-19 for UK Biobank participants with African ancestry, comparing number of *APOL1* variants carried to carriage of 0 variants. Adjusted for age, sex, body mass index, diagnosis of: chronic kidney disease, atrial fibrillation, depression, chronic obstructive pulmonary disease, dementia, type 2 diabetes, and UK Biobank genetic principal components 1-10. P values ≤0.05 are shown in bold.

| Risk Factor | N | Hospitalised<br>N (%) | Hospitalised<br>OR (95% CI) | Hospitalised<br>P | Death<br>N (%) | Death OR<br>(95% CI) | Death<br>P |
| --- | --- | --- | --- | --- | --- | --- | --- |
| 0 variants | 4,227 | 66 (1.6%) | - | - | 16 (0.4%) | - | - |
| 1 variant | 4,018 | 79 (2.0%) | 1.1 (0.7-1.5) | 0.8 | 19 (0.5%) | 1.2 (0.6-2.6) | 0.6 |
| 2 variants | 1,188 | 27 (2.3%) | 1.1 (0.7-1.8) | 0.6 | 12 (1.0%) | 2.7 (1.2-6.4) | **0.02** |

Studies of *APOL1*-associated kidney diseases have focused on the number of variant or risk alleles that an individual carries: carriage of two variant alleles (G1/G1, G2/G2, or G1/G2) is considered to be deleterious, while carriers of one variant allele (G0/G1 or G0/G2) have a risk of disease similar to those with no variants^5,6,7,8,9,10^.

Prior to its associations with non-communicable diseases, *APOL1* was identified as the trypanolytic protein, a pore-forming serum protein that inserts into membranes where it forms disruptive ion channels and lyses protozoan trypanosome parasites, protecting humans from infection by many trypanosome species^11^. The two subspecies of Trypanosoma brucei that infect humans have developed specific mechanisms for avoiding lysis by APOL1, either by binding, avoiding, or degrading the lytic protein^12^. Studies examining the role of *APOL1* in *T*.*b. gambiense* and *T*.*b. rhodesiense* infections have highlighted differences between G1 and G2. In T.b. rhodesiense, resistance to infection is only associated with the G2 variant^11^. Whereas in *T*.*b. gambiense* infection, carriage of G1 and G2 is associated with decreased and increased risk, respectively, of severe disease. Intriguingly, the G2-associated risk of severity of *gambiense* sleeping sickness is mitigated by the presence of G1 in the G1/G2 genotype^11^. It has been proposed that protection against human African trypanosomiasis might have driven selection for *APOL1* G1 and G2 in Africa^11^.

The membrane-damaging properties of APOL1 might be the common mechanism that explains both the destruction of parasites and damage to host tissues in other conditions^13^, however, several other pathways to disease have been proposed^14^. Recently, endothelial cell-specific expression of APOL1 risk alleles in mice was shown to induce mitophagy, leakage of mitochondrial DNA into the cytoplasm, and the activation the NLRP3 and STING innate immune pathways, which resulted in endothelial dysfunction^5^.

There is growing evidence that *APOL1* genotype also influences outcomes in COVID-19^15,16,17^. These investigations followed the precedent set by previous studies on *APOL1*-associated non-communicable diseases by considering the number of variant alleles carried rather than the complete genotype. Here, we use data from the UK Biobank to test the hypothesis that adverse outcomes in SARS-CoV-2 infection are associated with these variants, and present evidence that the association between carriage of two *APOL1* variants and adverse COVID-19 outcomes is driven primarily by one particular genotype: G1/G2.

## Methods

### Study design and participants

The UK Biobank is a large-scale biomedical database and research resource containing genetic, lifestyle, and health information from half a million participants from across the UK^18^. *APOL1* G1 and G2 are predominantly found in people with recent West African ancestry. UK Biobank participants with recent African ancestry were identified using previously reported^18^ principal components (PC1>100 and PC2 >0) and selected for inclusion in this study.

*APOL1* genotypes were obtained from the UK Biobank: the G1 (rs73885319) allele was genotyped using a custom Affymetrix array, and the genotype of the G2 (rs71785313) allele was obtained by imputation against the UK10K haplotype panel merged with the 1000 genomes phase 3 genotypes^18^.

Our cohort consisted of 10,190 individuals with African ancestry and accounted for all UK Biobank participants with G1/G1, G2/G2, or G1/G2 genotypes, 95% of participants who were G0/G1, and 94% of those who were G0/G2.

### COVID-19 diagnosis and outcome

Results of SARS-CoV-2 testing were listed in the UK Biobank tables ‘covid19_result_england’, ‘covid19_result_scotland’, or ‘covid19_result_wales’ of the UK Biobank’s Data Portal. Hospitalisations and deaths due to COVID-19 were coded ‘U071’ in the ‘hesin_diag’ and ‘death_cause’ tables, respectively. Deaths were attributed to COVID-19 if the term COVID-19 appeared on the death certificate. Among the cohort, 3,147 individuals had undergone a PCR test for SARS-CoV-2 with 824 testing positive; 172 had been hospitalised as a result of COVID-19, and 47 had died with COVID-19 listed as a cause.

### Covariates

Selection of covariates was based on previously described factors that were associated with poor COVID-19 outcomes (age, sex, chronic kidney disease, atrial fibrillation, depression, chronic obstructive pulmonary disorder, dementia, type 2 diabetes, body mass index, and Townsend deprivation index)^19,20,21^. UK Biobank principal components 1-10^18^ were also selected as covariates.

### Association analysis

The primary exposure variables were the number of *APOL1* variants carried and the six observed *APOL1* G1 and G2 genotypes (Supplementary Table 1). Firth’s Bias-Reduced Logistic Regression models were used to study the association of the exposures and the outcomes of interest. All logistic regression models passed the goodness of fit test by the Hosmer-Lemeshow test. All statistical tests were 2-sided, where a *P* = 0.05 or a 95% CI that did not contain unity were considered statistically significant for the primary outcome. All analyses were conducted using R statistical software (version 4.0.5.; R Foundation, Inc).

## Results

Of the 10,190 UK Biobank participants identified as having African ancestry using the previously reported principal component analysis^18^, 9,433 had complete *APOL1* genotype data and were included in this study.

Chi-squared tests confirmed that carriage of either one or two *APOL1* variant alleles was not associated with SARS-CoV-2 testing (one variant, p=0.09; two variants p=0.13) or positivity (one variant, p=0.52; two variants p=0.94). This indicated that carriage of *APOL1* variants is not associated with the likelihood of undergoing a PCR test or receiving a positive PCR test result.

We then tested the hypothesis that carriage of one (G0/G1 or G0/G2) or two (G1/G1, G1/G2, or G2/G2) variants are associated with severe SARS-CoV-2 infection outcome (either hospitalization or death) using Firth’s Bias-Reduced Logistic Regression (Table 1). The first 10 UK Biobank principal components were included as covariates along with previously-associated COVID-19 risk factors (age, sex, chronic kidney disease, atrial fibrillation, depression, chronic obstructive pulmonary disorder, dementia, type 2 diabetes, body mass index, and Townsend deprivation index)^19,20,21^. There was no association with carriage of one or two APOL1 variants and hospitalisation but there was an association (OR= 2.7, 95% CI: 1.2-6.4, p=0.02) between carriage of two variants and death due to COVID-19 (Table 1).

Having observed an association with carriage of two *APOL1* variants, we then explored this association in more detail by examining all genotype combinations that have been detected in populations with recent African ancestry. No association was detected between hospitalisation or death with G0/G1, G0/G2, G1/G1, or G2/G2 when compared with G0/G0. However, a strong association was detected for the G1/G2 genotype with hospitalisation (OR = 2.1, 95% CI: 1.1-3.8, p = 0.02) as well as death (OR = 5.9, 95% CI: 2.2-15.3, p = 0.0007) (Table 2). This indicates that the association between G1/G2 (carried by 343 individuals in our cohort (3.6%)) and COVID-19 outcome is primarily responsible for the positive association with carriage of two variants described above.

**Table 2:** Risk of hospitalisation or death due to COVID-19 for UK Biobank participants with African ancestry, comparing different *APOL1* genotypes to G0/G0. Adjusted for age, sex, body mass index, diagnosis of: chronic kidney disease, atrial fibrillation, depression, chronic obstructive pulmonary disease, dementia, type 2 diabetes, and UK Biobank genetic principal components 1-10. P values ≤0.05 are shown in bold.

| Risk Factor | N | Hospitalised<br>N (%) | Hospitalised<br>OR (95% CI) | Hospitalised<br>P | Death<br>N (%) | Death OR<br>(95% CI) | Death<br>P |
| --- | --- | --- | --- | --- | --- | --- | --- |
| G0/G0 | 4,227 | 66 (1.6%) | - | - | 16 (0.4%) | - | - |
| G0/G1 | 2,600 | 52 (2.0%) | 1.1 (0.7-1.6) | 0.7 | 13 (0.5%) | 1.4 (0.6-3.2) | 0.4 |
| G0/G2 | 1,418 | 27 (1.9%) | 1.0 (0.6-1.6) | 1.0 | 6 (0.4%) | 1.0 (0.4-2.7) | 0.9 |
| G1/G1 | 687 | 10 (1.5%) | 0.8 (0.4-1.5) | 0.5 | 3 (0.4%) | 1.5 (0.4-4.6) | 0.5 |
| G1/G2 | 343 | 15 (4.4%) | 2.1 (1.1-3.8) | **0.02** | 8 (2.3%) | 5.9 (2.2-15.3) | **0.0007** |
| G2/G2 | 158 | 2 (1.3%) | 0.8 (0.2-2.3) | 0.7 | 1 (0.6%) | 2.4 (0.3-10.7) | 0.4 |

## Discussion

There is growing evidence that carriage of two high-risk *APOL1* variants is associated with poor outcomes in COVID-19. In African Americans infected with SARS-CoV-2, carriage of two *APOL1* variants is associated with collapsing glomerulopathy^15^, acute kidney injury (AKI), persistent AKI, and requirement for kidney replacement therapy^16^. Among African Americans hospitalised with COVID-19, carriage of two high-risk APOL1 variants has been associated with increased AKI severity and death^17^. These investigations followed the precedent set by previous studies on *APOL1*-associated non-communicable diseases by considering the number of variant alleles carried rather than the individual genotypes.

UK Biobank participants of African ancestry who have been hospitalised and/or died from COVID-19 represent a relatively small dataset, and verification in a larger cohort is required. However, our observations support extending the risk factors associated with poor COVID-19 outcomes in people of African ancestry to *APOL1* genotype. Although they provide further evidence of a relationship between carriage of two *APOL1* variants and adverse COVID-19 outcomes, they also indicate that the deleterious effects are driven primarily by the G1/G2 genotype. G1/G2 is a multi-locus genotype, possibly explaining why it was not previously detected in genome wide association studies for COVID-19^22,23^. Differential function of *APOL1* G1 and G2 has previously been highlighted in human African trypanosomiasis^11^, and these differences might be relevant to mechanisms leading to the varied inflammatory-mediated diseases and need to be further explored.

Evidence of specifically the G1/G2 genotype being associated with deleterious outcomes in other diseases is limited. A previous study of HIV-associated nephropathy (HIVAN) patients in South Africa reported that the G1/G2 genotype accounted for 50% of the 38 cases, far more than would be predicted from the local allele frequencies (estimated to be less than 6% for each allele)^10^. The work presented here supports a change in the way that association studies should be approached for *APOL1*: examining associations with individual genotypes rather than associations with carriage of two variants, which dominates the literature^5,6,7,8,9,10^.

This association between the G1/G2 genotype and adverse COVID-19 outcomes could have important implications, both at the individual and population level, by identifying those at higher risk of severe COVID-19 who would benefit from early vaccination and treatment. This is especially relevant to geographical regions where *APOL1* G1/G2 genotypes are common such as West and Central Africa, which have reported allele frequencies of up to 49% for G1 and 21% for G2^6,11^. Similarly, the African diaspora, which accounts for 140 million individuals world-wide represents a population with significant enhanced risk of adverse COVID-19 outcomes.

## Data Availability

This research has been conducted using the UK Biobank Resource under application number 66821. All bona fide researchers can apply to use the UK Biobank resource for health related research that is in the public interest.

https://www.ukbiobank.ac.uk

## Approval

Access to the UK Biobank data was granted for this work under UK Biobank application number 66821.

