## Supplementary Table 1 for "Adverse outcomes in SARS-CoV-2 infections are associated with a combination of variant genotypes at two loci in the *APOL1* gene: a UK Biobank study"

**Supplementary data**

| Genotype Name | Haplotypes  G1 locus G2 locus | | Number of variants |
| --- | --- | --- | --- |
| G0/  G0 | AT  AT | TTATAA  TTATAA | 0 |
| G0/  G1 | AT  GG | TTATAA  TTATAA | 1 |
| G0/  G2 | AT  AT | TTATAA  ------ | 1 |
| G1/  G1 | GG  GG | TTATAA  TTATAA | 2 |
| G1/  G2 | GG  AT | TTATAA  ------ | 2 |
| G2/  G2 | AT  AT | ------  ------ | 2 |
| G1/  G1+G2* | GG  GG | TTATAA  ------ | 3 |
| G2/  G1+G2* | AT  GG | ------  ------ | 3 |
| G1+G2/  G2+G1* | GG  GG | ------  ------ | 4 |

**Supplementary Table 1:** Possible haplotype arrangements at the APOL1 G1 and G2 loci. G1 is rs73885319 (A/G) and rs60910145 (T/G) which are 128bp apart and in almost perfect linkage so that only the AT and GG haplotypes are nearly always observed. G2 (rs71785313) TTATAA/------ and is also linked to G1 so that only four haplotypes are observed (ATTTATAA, AT------ , GGTTATAA, GG------) but no recombinants between the alternate alleles. Genotypes marked with an asterisk are theoretically possible but have not been observed because they require carriage of G1 and G2 on the same haplotype.
